# The Analgesic Efficacy of Different Techniques Surrounding Regional Anesthesia of the Lumbar Plexus and its Terminal Branches for Hip Fracture Surgeries

**DOI:** 10.1101/2022.06.22.22276758

**Authors:** Abnoos Mosleh-Shirazi, Brian O’Donnell

## Abstract

**Background:** Research is limited in comparing the analgesic efficacy of the various types of blocks with one another for hip fracture surgeries. Due to the rapid pace in the development of these new techniques in blocking the lumbar plexus and its terminal branches, uncertainty exists in literature and in practice regarding the definition and efficacy of one technique in comparison to another.

**Objectives:** (1) To write a narrative description of regional anesthesia approaches to the lumbar plexus and associated terminal branches; (2) To do a systematic review and meta-analysis of published articles regarding the analgesic efficacy of regional anesthesia in the context of hip fracture and hip fracture surgery.

**Questions:** (1) Does regional anesthesia of the lumbar plexus and its terminal branches enhance analgesic outcomes following hip fracture and hip fracture surgery? (2) Does the evidence point toward one techniques superiority over another? (3) Does evidence show a necessity for a nerve block over the use of opioid analgesics?

**Search methods:** Six databases: EMBASE, PUBMED, SCOPUS, EBSCO (CINAHL and MEDLINE), WEB OF SCIENCE, COCHRANE LIBRARY were searched on October 12th, 2020.

**Search criteria:** Studies were selected based on inclusion of: Study Design: Prospective Randomized Controlled Trials (RCT), Population: Adults (18+ years) undergoing hip fracture surgery, Intervention: FNB, FICB, PCB and/or PENG block, Comparison: Another intervention of interest, Placebo, Non-intervention, Systemic analgesics (Opioids, NSAIDs, Paracetamol), Outcome: Analgesic efficacy (Pain scores measured by Numeric Pain Rating Scale (NRS) or Visual Analogue Scale (VAS)). Studies were excluded if: Unavailable in full-text, non-human studies, Not RCT, Surgery unrelated to hip fracture.

**Data collection and analysis:** Two reviewers extracted all relevant data from the full text versions of eligible studies using a predefined data extraction form. Study characteristics included: author, publication year, study design, sample size, inclusion and exclusion criteria, type of intervention and control, statistical analysis, outcome data, and authors’ main conclusions.

Risk of bias in individual studies assessed by two reviewers based on criteria adapted from the Cochrane ‘Risk of Bias’ assessment tool. High-risk studies were excluded.

**Main results:** 1. FICB vs Opioid: pain scores at rest at 24h were lower in the FICB group (-0.79 [-1.34, - 0.24], P= 0.005). Pain scores on movement at 12h were lower in the FICB group (-1.91 [-2.5, -1.3], P<0.00001). No difference between groups in other times. 2. FNB vs Opioid: Initial pain scores at rest were lower in FNB (-0.58 [-0.104, -0.12], P=0.01). 3. FICB vs FNB: No difference between groups at rest. Pain scores on movement: initial scores following block, and at 24 hours were lower in the FNB group (initial: 0.53 [0.21, 0.86], P=0.001, 24 h: 0.61 [0.29, 0.94], P=0.0002, results not estimable for 12h (not enough data)).

**Authors’ conclusions:** Both femoral nerve block and fascia iliaca compartment block enhance analgesic outcomes following hip fracture and hip fracture surgery, superior to the use of systemic analgesics such as opioids. FNB may be more efficacious at reducing pain following hip fracture surgery when compared to FICB.

## INTRODUCTION

### Description of the condition

Hip fractures are a common occurrence in elderly patients with multiple comorbidities such as hypertension, diabetes mellitus, cardiovascular disease, and more. Consequently, elderly patients are usually on long-term medications, which result in hemodynamic instability that complicates the perioperative management of hip-fracture patients. Unfortunately, there is high morbidity and mortality among these patients, and they are often undertreated for pain. Due to physiological frailty, medical comorbidities and cognitive impairment interfering with pain assessment and treatment, pain management in the elderly is challenging,[1].

### Description of the intervention

Opioids, along with other multimodal techniques, are commonly used to address pain. However due to altered pharmacodynamics and coexisting medical conditions, elderly patients are vulnerable to the side effects of opioids such as respiratory depression[2].

The use of peripheral nerve blockade for pain management may resolve some of the issues surrounding the use of opioid analgesics in the elderly. Targeted somatic nerve block via regional anesthesia provides rapid-onset, site specific analgesia while preventing dispensable sympathetic block even in CVS compromised patients[3].

In 1884, the first clinical locoregional anesthetic technique was introduced,[4] followed by the first central neuraxis block just 5 years later. After another 7 decades, the first proximal lower extremity peripheral nerve block was described in 1973 called the “3-in-1 block,”[4]. Since then, many techniques for regional anesthesia of the lumbar plexus and its terminal branches have evolved; including psoas compartment block (PCB) (posterior lumbar plexus block), Fascia iliaca compartment block (FICB), 3-in-1 femoral nerve block (FNB) (anterior lumbar plexus block), PENG (pericapsular nerve group) block, etc. These techniques are frequently used for hip fracture patients due to the opioid-sparing effects,[5].

### How the intervention might work

The lumbar plexus is formed by lumbar nerves L1-L4 and is the origin of the ilioinguinal and iliohypogastric nerves, as well as the 4 major nerves that supply the lower limb: femoral, lateral femoral cutaneous, obturator, and genitofemoral. The plexus is formed lateral to the intervertebral foramina and passes through psoas major. The nerves of the lumbar plexus pass in front of the hip joint and mainly support the anterior part of the thigh[7].

Regional anaesthesia provides analgesia by pharmacologic disruption of nociceptive transmission, neuroanatomy of the injury, anatomical target sites. Analgesia of large parts of the leg can be achieved by blocking the lumbar plexus and its associated terminal branches. Thus, it is principally used for post-operative analgesia for hip surgeries[8] and provide opioid-sparing effects[9].

A timeline and description of regional anaesthesia approaches to the lumbar plexus and associated terminal branches is provided in Appendix A.

### Why it is important to do this review

Early surgery within 48 hours of the hip fracture has shown to decrease complication and mortality rates[6]. This gives an opportunity for anesthesiologists to offer regional analgesia for effective pain control. However, due to the rapid pace in the development of these new techniques in blocking the lumbar plexus and its terminal branches, confusion exists in literature and in practice regarding the definition and efficacy of one technique in comparison to another.

### Objectives

(1) To write a narrative description of regional anesthesia approaches to the lumbar plexus and associated terminal branches; (2) To do a systematic review and meta-analysis of published articles regarding the analgesic efficacy of regional anesthesia in the context of hip fracture and hip fracture surgery, to answer the following questions: (1) Does regional anesthesia of the lumbar plexus and its terminal branches enhance analgesic outcomes following hip fracture and hip fracture surgery? (2) Does the evidence point toward one techniques superiority over another? (3) Does evidence show a necessity for a nerve block over the use of opioid analgesics?

## METHODS

### Criteria for considering studies for this review

#### Types of studies

Only prospective RCTs were included in this study, including no blinding, single-blinded and double-blinded studies.

#### Types of participants

Adults (18+ years) undergoing hip fracture surgery.

Outcome: Analgesic efficacy (Pain scores measured by Visual analogue scale (VAS) or Numerical rating scale (NRS)). Studies were excluded if: Unavailable in full-text, non-human studies, Not RCT, Surgery unrelated to hip fracture.

#### Types of interventions

FNB, FICB, PCB and/or PENG block compared to another intervention of interest, placebo, non-intervention, or systemic analgesics (opioids, NSAIDs, paracetamol). Any dose or form of local anesthetic to accompany technique (including continuous catheter infusion or single dose) were included.

#### Types of outcome measures

##### Primary outcomes

Analgesic efficacy of the intervention using VAS or NRS scales.

#### Secondary outcomes

Other outcomes were accounted for but not included in the meta-analysis, such as: pre-operative and/or post-operative need for analgesia, time to first request for additional analgesia, adverse effects, allergic reactions, length of hospital stay, mortality.

### Search methods for identification of studies

#### Electronic searches

Both electronic and hand-searching techniques were used to identify studies. Six databases: EMBASE, PUBMED, SCOPUS, EBSCO (CINAHL and MEDLINE), WEB OF SCIENCE, COCHRANE LIBRARY were searched on October 12th, 2020.

Searches were limited based on inclusion criteria:

Type of block: Femoral nerve block, Fascia iliaca compartment block, Psoas compartment block, Lumbar plexus block, Pericapsular nerve group PENG block

Type of study: Randomized controlled trial, Human study, Available in English

Age: Adult (18+)

Search words for each database are summarized in Appendix B.

### Searching other resources

Manual searches were then conducted for all eligible articles via their reference list and related articles suggested by PubMed.

### Data collection and analysis

Two reviewers extracted all relevant data from the full text versions of eligible studies using a predefined data extraction form. Study characteristics included: author, publication year, study design, sample size, inclusion and exclusion criteria, type of intervention and control, statistical analysis, outcome data, and authors’ main conclusions.

#### Selection of studies

The abstract of each study was reviewed for eligibility by one reviewer (A.M.S). Potential studies underwent full text review by the same author. Studies were excluded if they did not meet the inclusion criteria. The inclusion and exclusion criteria are outlined in Appendix C.

#### Assessment of risk of bias in included studies

Risk of bias in individual studies assessed by two reviewers based on criteria adapted from the Cochrane ‘Risk of Bias’ assessment tool (RoB 2). A study was rated overall as Low risk, Some concerns, or High risk for bias based on five domains: (1) Randomisation process, (2) Deviations from intended interventions, (3) Missing outcome data, (4) Measurement of the outcome, (5) Selection of the reported result. High risk of bias determined by important imbalances at baseline, improper randomisation, failure of blinding of outcome assessors and significant (>15%) loss to follow-up. High-risk studies were excluded.

#### Unit of analysis issues

The Hozo equation (Hozo) was used to derive mean and standard deviation from median and inter-quantile ranges. For sample sizes greater than 25, the sample’s median was set to equal to mean as recommended by Hozo et al. Also, for sample sizes of 15 through 70, range was divided by 4 as the best estimate of the sample’s standard deviation as recommended by Hozo et al. The 11-point NRS, and 100-mm VAS scores divided by 10 were considered equal to the 10-cm VAS scale. Point estimates and 95% confidence intervals (CIs) of individual included studies and results were shown via forest plots. Data analyses abided by the guidelines set out by the Cochrane Collaboration regarding statistical methods. two-tailed P-values <0.05 were considered significant in all instances. Relative risks and the standardised mean difference (SMD) for continuous outcomes were also calculated.

#### Dealing with missing data

Authors of included studies were individually contacted for missing data.

#### Assessment of heterogeneity

Chi-squared statistic was used to assess heterogeneity, where values >50% are consistent with large heterogeneity, and using heterogeneity P-value, where values <10% are consistent with large heterogeneity.

#### Assessment of reporting biases

Funnel plots were conducted to detect publication bias.

#### Subgroup analysis and investigation of heterogeneity

Since heterogeneity was expected across studies, an inverse variance random-effects model was used to evaluate outcomes. Heterogeneous control interventions and data collection points were investigated via subgroup analyses. Intervention types were pooled into the following subgroups; (1) Fascia-iliaca compartment block vs. intra-articular hip injection (2) Fascia-iliaca compartment block vs. Placebo (sham block) (3) Fascia iliaca compartment block vs. Femoral nerve block (4) Fascia iliaca compartment block vs. opioid or other analgesic (5) Femoral nerve block vs. Placebo (sham block) (6) Femoral nerve block vs. opioid or other analgesic.

#### Sensitivity analysis

All analyses were conducted using Review Manager (RevMan; version 5.4).

A flow chart demonstrating an overview of the research protocol and study selection process can be found in Appendix D.

## RESULTS

### Description of the studies

#### Results of the search

The database search yielded 439 studies. A total of 316 abstracts were screened after removal of duplicates. After screening, 70 full text articles were assessed for eligibility.

#### Included studies

In total, twenty-five studies met the inclusion criteria[12, 14, 16, 20, 23, 27, 28, 33, 34, 38, 47, 51, 59, 60, 62, 65, 57, 66, 74, 77, 80, 82, 83, 85, 86]. Included studies date from 2007 to 2019.

Five articles were removed due to high risk of bias[27, 33, 51, 74, 80], leaving twenty studies for the meta-analysis. The twenty studies yielded 1871 patients in total, with 937 total patients in the intervention groups, and 934 total patients in the control groups.

#### Excluded studies

Eight studies were removed because the research was not yet complete[17, 18, 53-58], 20 studies were removed because they were irrelevant to hip fracture[10, 13, 15, 22, 24, 26, 29-32, 39, 41, 43, 45, 63, 68, 69-71, 73], six studies were removed because their outcomes did not focus on analgesic efficacy[11, 40, 52, 64, 78, 79], two studies were unavailable in full text (19, 75), and lastly, 9 articles were removed due to inappropriate study design[25, 35, 36, 42, 44, 49, 50, 67, 84]. The reference lists of all articles examined by full text and similar reviews were hand searched, but yielded no additional articles.

#### Study characteristics

Table 1 presents highlighted study features. All twenty studies were randomised controlled trials. Seven studies compared FICB with opioids[28, 47, 60, 62, 82, 83, 85]. Seven studies compared FNB with opioids[14, 16, 34, 38, 66, 72, 77]. Five studies compared FNB with FICB [20, 23, 59, 65, 86]. One study compared FICB with intra-articular hip injection[12]. See Table 1.

**Table 1.**
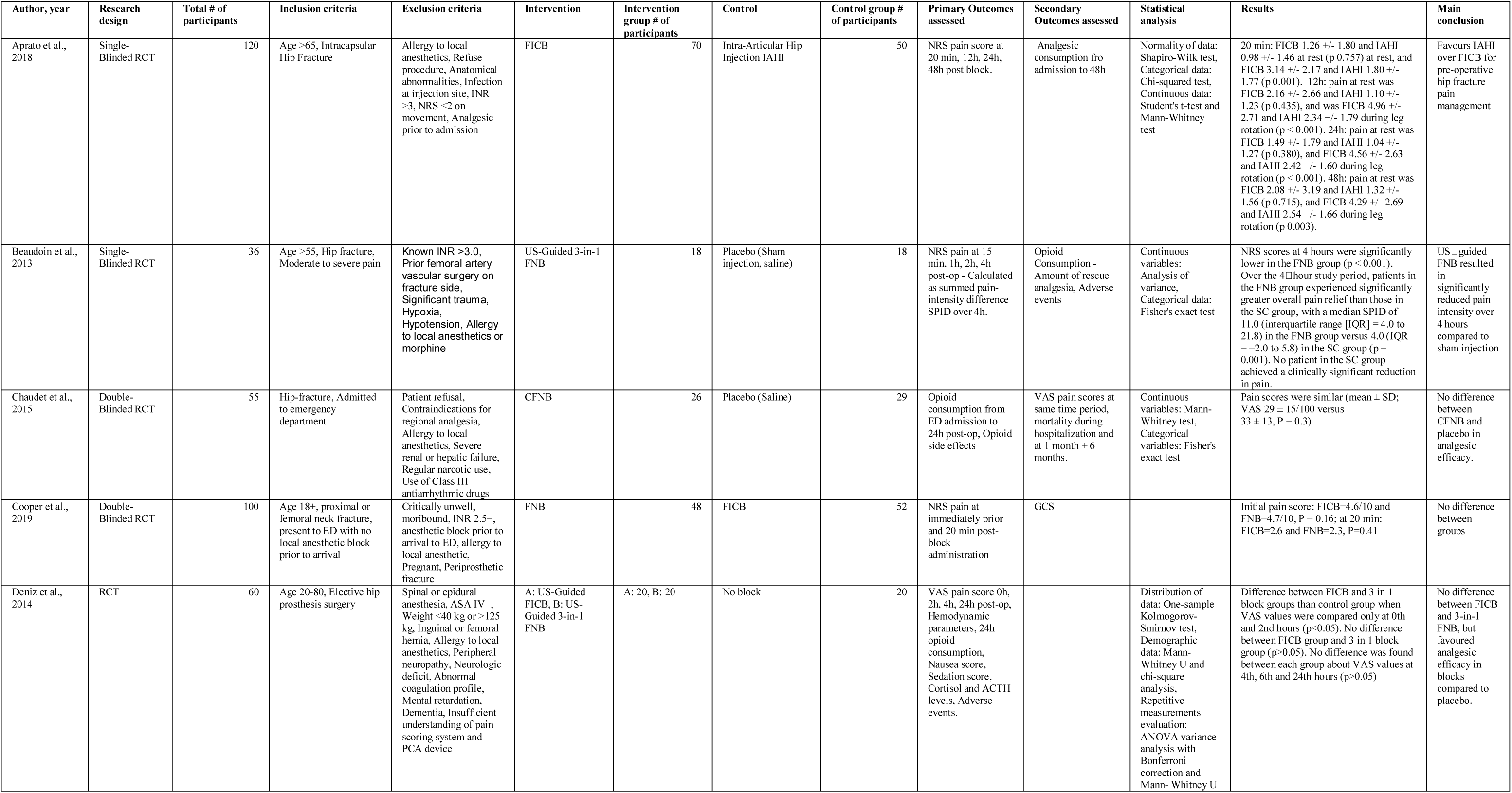
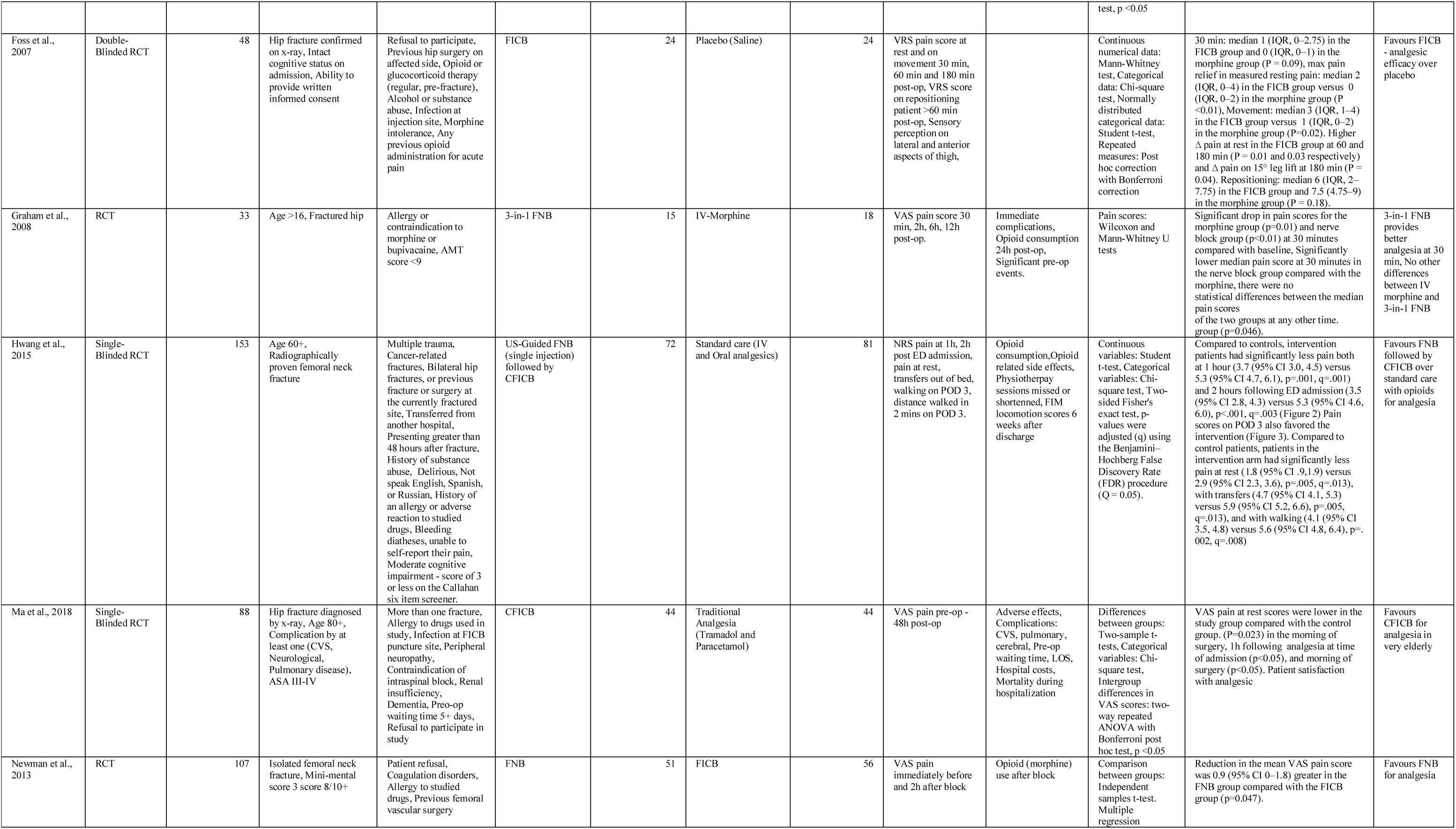
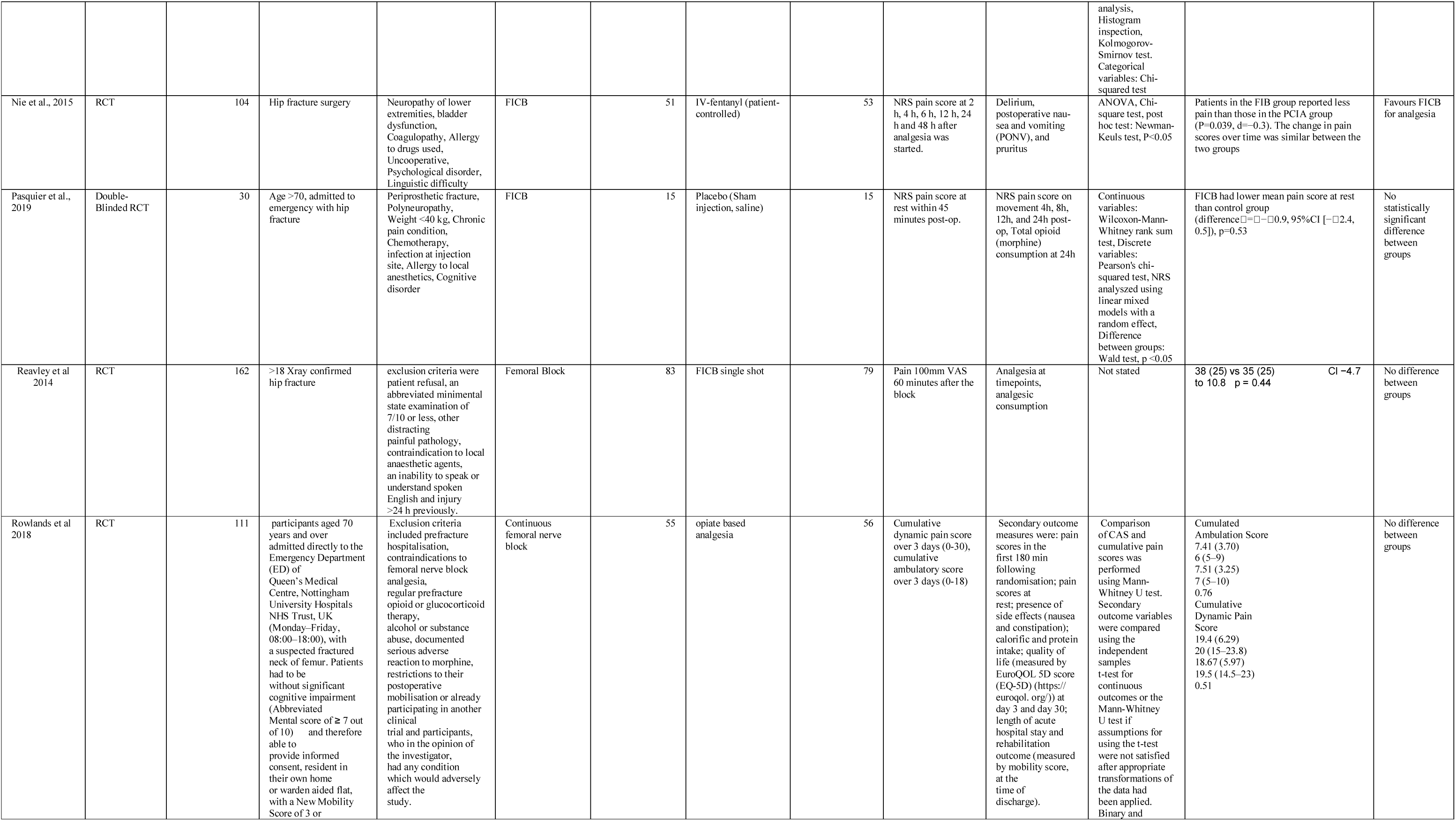
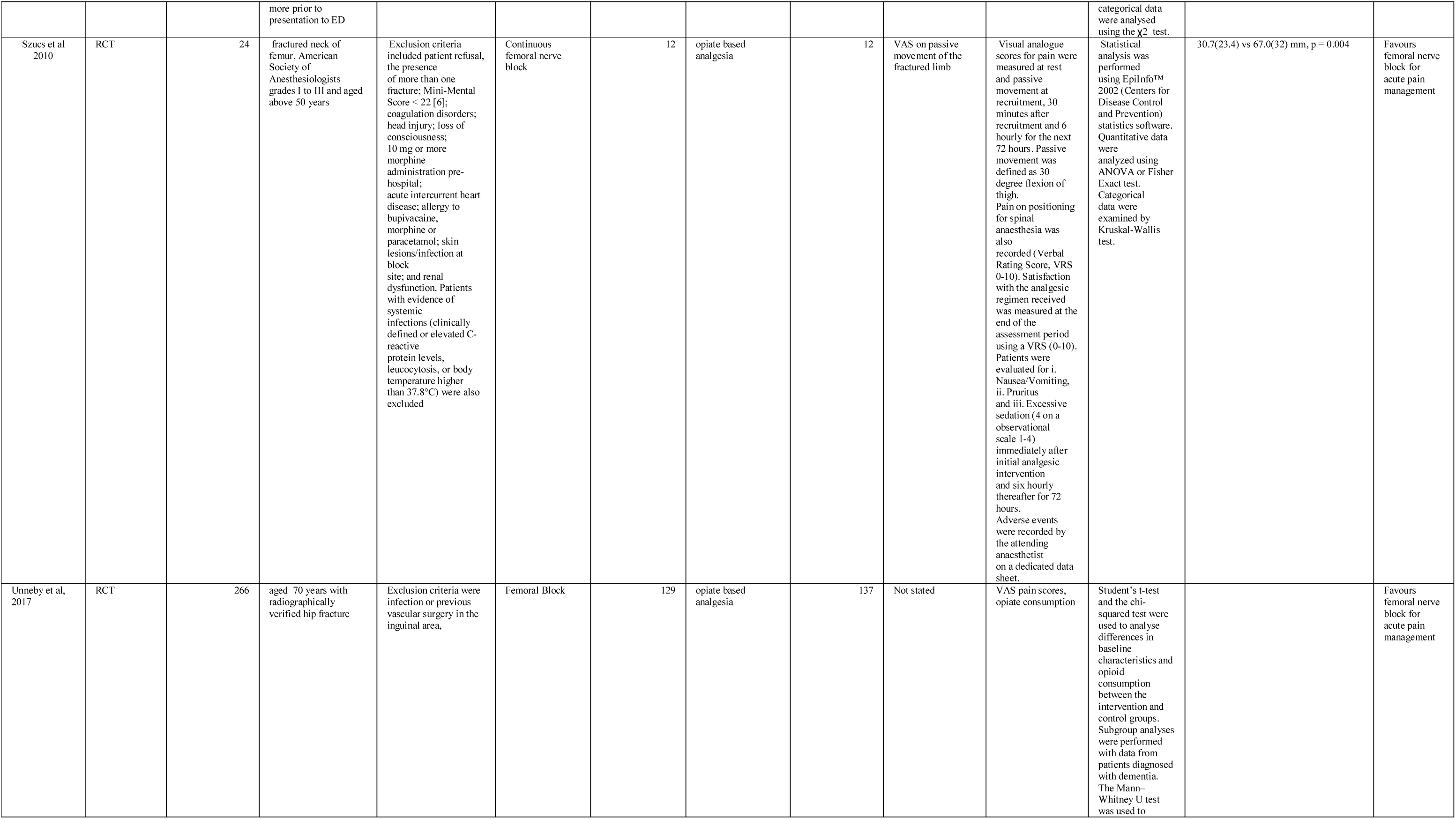
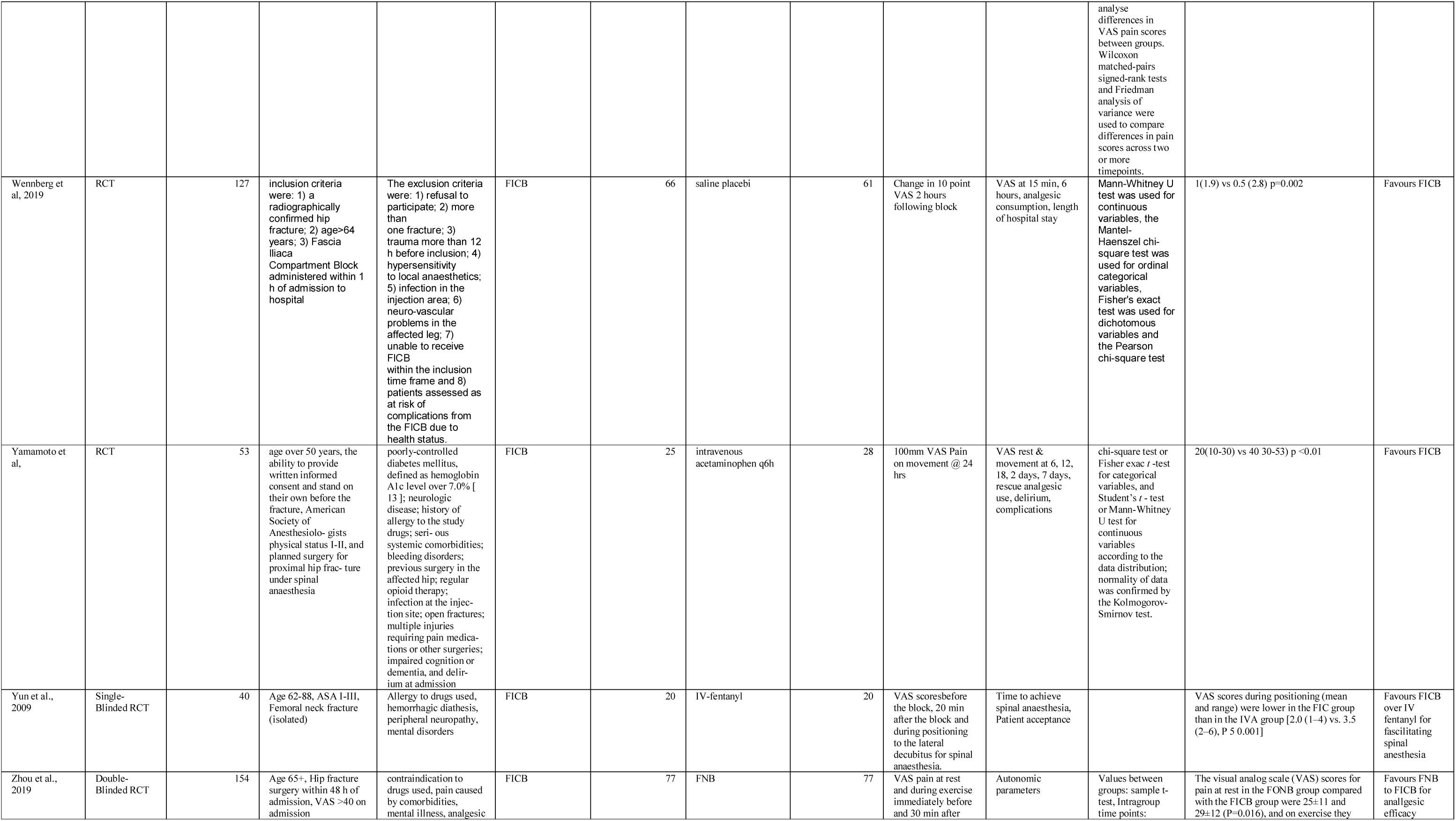
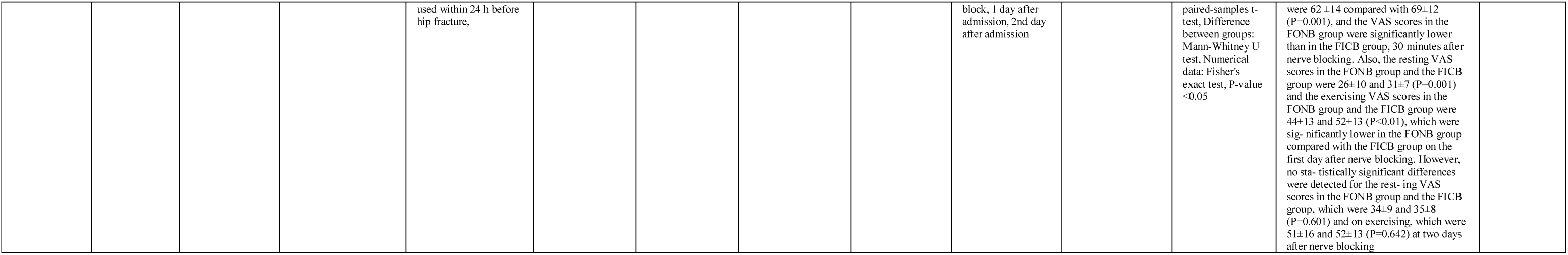
Study characteristics for all included studies in this review. (FICB, fascia iliaca compartment block; FNB, femoral nerve block; NRS, numeric rating scale; VAS, visual analogue scale; RCT, randomized controlled trial; US, ultrasound).

#### Risk of bias in included studies

Based on criteria adapted from the Cochrane ‘Risk of Bias’ assessment tool, nine of the included randomised controlled trials[12, 16, 38, 47, 62, 65, 82, 85, 86] were rated with overall low risk of bias, eleven studies had some concerns according to RoB 2[14, 20, 23, 28, 34, 59, 60, 66, 72, 77, 83], and five studies were rated as high risk of bias[27, 33, 51, 74, 80]. These findings are summarized in Table 2. Of the papers with some concern for risk of bias, five had issues with the randomisation process[14, 20, 23, 28, 77], two had deviations from the intended interventions[14, 57], eight had issues with measurement of the outcome[23, 34, 59, 60, 66, 72, 77, 83]. Of the papers with high risk of bias, three had issues with; the (i) randomization process, had (ii) deviations from the intended interventions, and had issues with the (iii) outcome measurement[27, 74, 80], one had issues with the randomization process and measurement of the outcome[33], and one had deviations from the intended interventions as well as issues with the measurement of outcome[51]. None of the papers had missing outcome data, loss to follow-up >15%. or issues with selection of the reported result. The five papers with high risk of bias according to the RoB 2 tool were excluded from the meta-analysis. Thus, twenty articles were included in the meta-analysis.

**Table 2.**
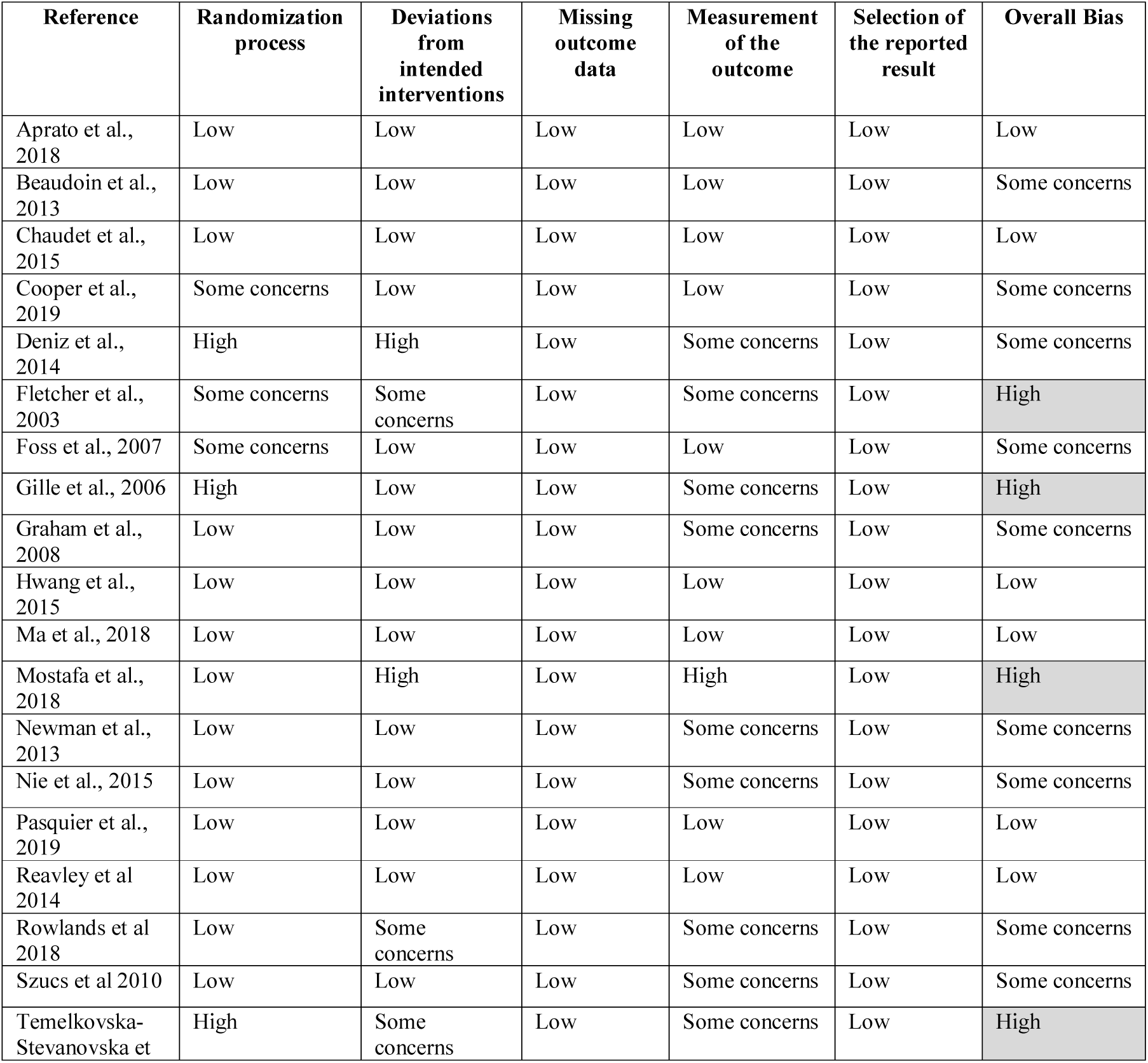
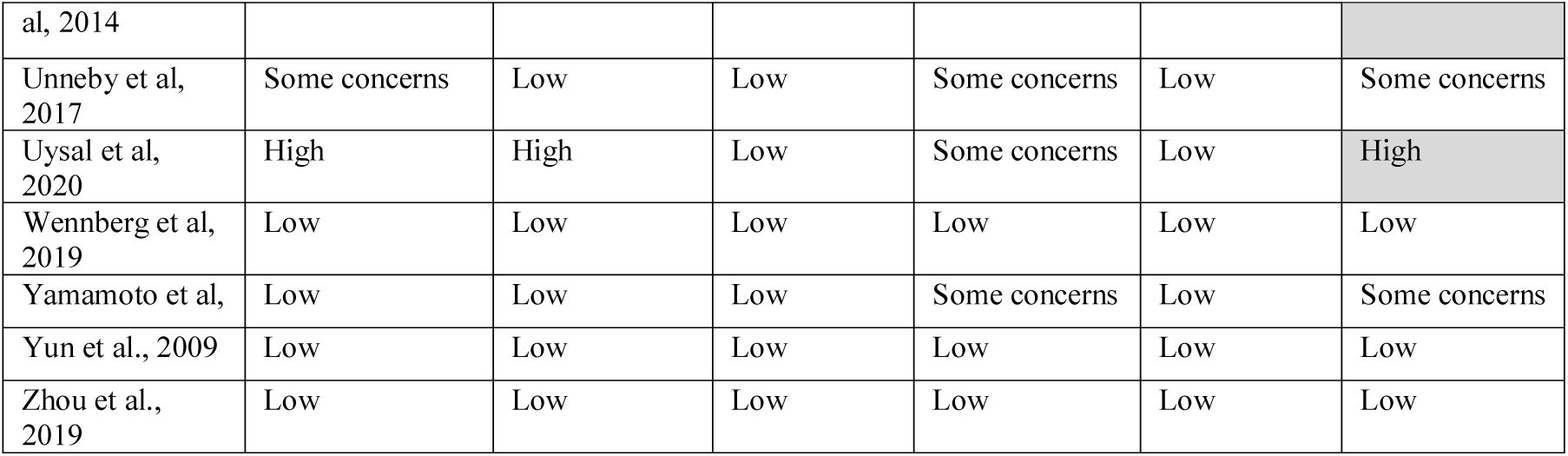
Risk of bias within studies based on criteria adapted from the Cochrane ’Risk of Bias’ assessment tool.

#### Results of individual studies

In the meta-analysis of the seven studies comparing the analgesic efficacy of FICB compared with Opioids, there was a statistically significant difference between the groups at rest at 24 hours and on movement at 12 hours as shown in Table 3. Data for pain scores at rest at 24 hours in this group were derived from three studies out of the seven as shown in Figure 1. Compared with opioids at rest at 24 hours, FICB had a greater analgesic effect with an SMD (standardized mean difference) of -0.79 (95% CI; -1.34, -0.24) (P = 0.005) (Z = 2.83) (heterogeneity: tau^2^ = 0.09, Chi^2^ = 3.23, df = 2, P = 0.20, I^2^ = 38%). Data for pain scores on movement at 12 hours in this group were derived from two studies out of the seven as shown in Figure 2. Compared with opioids on movement at 12 hours, FICB had a greater analgesic effect with an SMD (standardized mean difference) of -1.91 (95% CI; -2.52, -1.30) (P < 0.00001) (Z = 6.18) (heterogeneity: tau^2^ = 0.00, Chi^2^ = 1.00, df = 1, P = 0.32, I^2^ = 0%).

**Figure 1.**
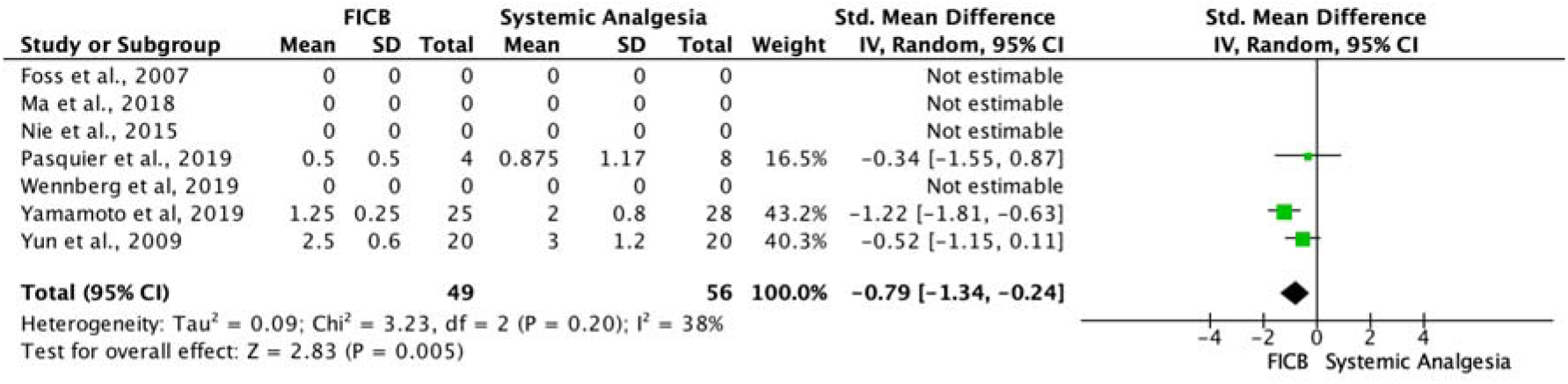
Forest plot of comparison: FICB vs. Opioids. Outcome: pain scores at rest at 24 hours.

**Figure 2.**
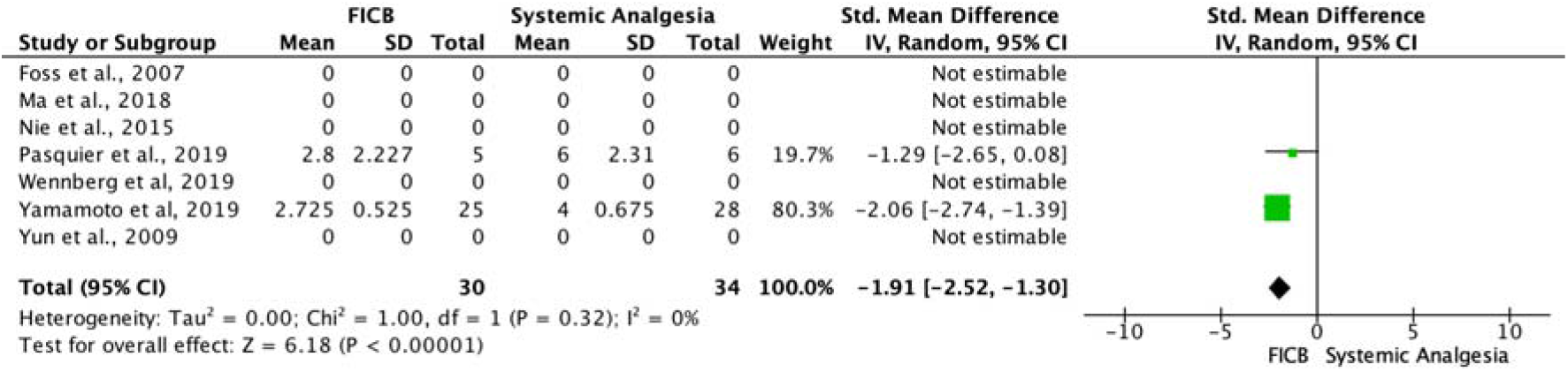
Forest plot of comparison: FICB vs. Opioids. Outcome: pain scores on movement at 12 hours.

**Table 3.** Fascia iliaca compartment block vs. Opioids

| Outcome or Subgroup | Number of Studies | Participants | Statistical Method | Effect Estimate |
| --- | --- | --- | --- | --- |
| Initial Pain Scores at | 7 | 285 | Std. Mean Difference | 0.05 [-0.78, 0.88] |
| Rest Following Block |  |  | (IV, Random, 95% CI) |  |
| Pain Scores at Rest at 12 Hours | 7 | 67 | Std. Mean Difference (IV, Random, 95% CI) | -1.29 [-2.93, 0.35] |
| Pain Scores at Rest at 24 Hours | 7 | 105 | Std. Mean Difference (IV, Random, 95% CI) | -0.79 [-1.34, -0.24] |
| Initial Pain Scores on Movement Following Block | 7 | 245 | Std. Mean Difference (IV, Random, 95% CI) | -0.87 [-3.12, 1.37] |
| Pain Scores on Movement at 12 Hours | 7 | 64 | Std. Mean Difference (IV, Random, 95% CI) | -1.91 [-2.52, -1.30] |
| Pain Scores on Movement at 24 Hours | 7 | 64 | Std. Mean Difference (IV, Random, 95% CI) | -2.01 [-5.27, 1.26] |

In the meta-analysis of the seven studies comparing the analgesic efficacy of FNB compared with Opioids, there was a statistically significant difference between the groups initially at rest following the block as shown in Table 4. Data for pain scores at rest following the block in this group were derived from five studies out of the seven as shown in Figure 3. Compared with opioids at rest following the block, FICB had a greater analgesic effect with an SMD (standardized mean difference) of -0.58 (95% CI; -1.04, -0.12) (P = 0.01) (Z = 2.49) (heterogeneity: tau^2^ = 0.19, Chi^2^ = 16.46, df = 4, P = 0.002, I^2^ = 76%).

**Figure 3.**
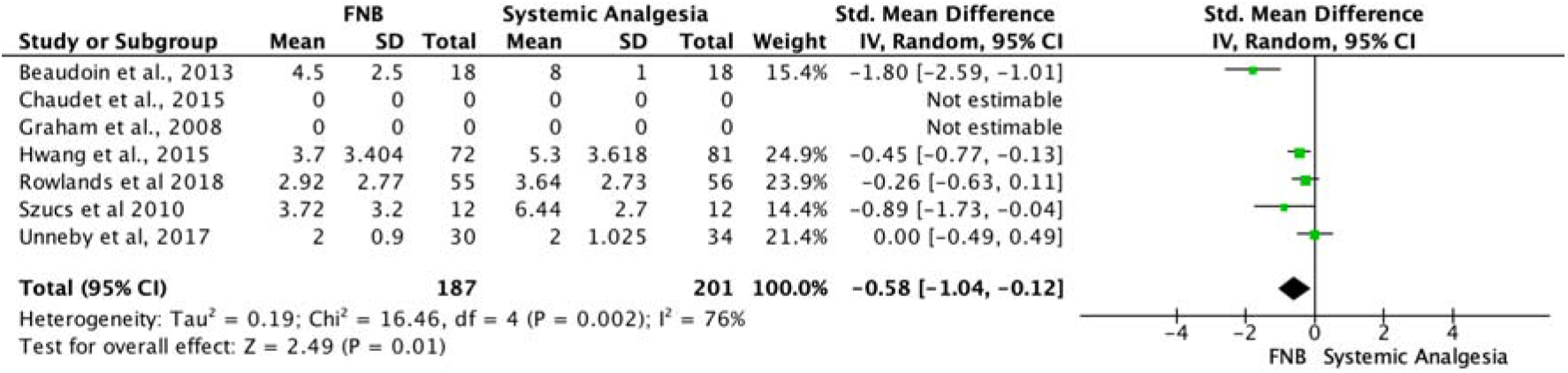
Forest plot comparison: FNB vs. Opioids. Outcome: initial pain scores at rest following block.

**Table 4.** Femoral nerve block vs. Opioids

| Outcome or Subgroup | Number of Studies | Participants | Statistical Method | Effect Estimate |
| --- | --- | --- | --- | --- |
| Initial Pain Scores at Rest Following Block | 7 | 388 | Std. Mean Difference (IV, Random, 95% CI) | -0.58 [-1.04, -0.12] |
| Pain Scores at Rest at 12 Hours | 7 | 64 | Std. Mean Difference (IV, Random, 95% CI) | -0.47 [-0.97, 0.03] |
| Pain Scores at Rest at 24 Hours | 7 | 55 | Std. Mean Difference (IV, Random, 95% CI) | -0.28 [-0.81, 0.25] |
| Initial Pain Scores on Movement Following Block | 7 | 111 | Std. Mean Difference (IV, Random, 95% CI) | -0.37 [-0.74, 0.01] |
| Pain Scores on Movement at 12 Hours | 7 | 0 | Std. Mean Difference (IV, Random, 95% CI) | Not estimable |
| Pain Scores on Movement at 24 Hours | 7 | 0 | Std. Mean Difference (IV, Random, 95% CI) | Not estimable |

In the meta-analysis of the five studies comparing the analgesic efficacy of FICB compared with FNB, there was a statistically significant difference between the groups on movement initially following the block and on movement at 24 hours as shown in Table 5. Data for pain scores on movement initially following the block in this group were derived from one study out of the five as shown in Figure 4. Compared with FICB on movement initially following the block, FNB had a greater analgesic effect with an SMD (standardized mean difference) of 0.53 (95% CI; 0.21, 0.86) (P = 0.001) (Z = 3.26) (heterogeneity: not applicable). Data for pain scores on movement at 24 hours in this group were derived from one study out of the five as shown in Figure 5. Compared with FICB on movement at 24 hours, FNB had a greater analgesic effect with an SMD (standardized mean difference) of 0.61 (95% CI; 0.29, 0.94) (P < 0.002) (Z = 3.71) (heterogeneity: not applicable).

**Figure 4.**
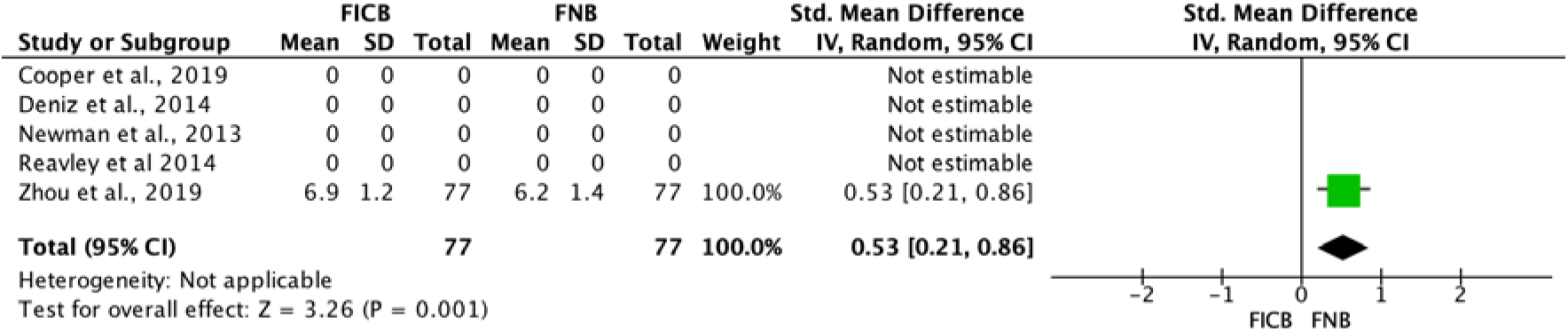
Forest plot comparison: FICB vs. FNB. Outcome: initial pain scores on movement following block.

**Figure 5.**
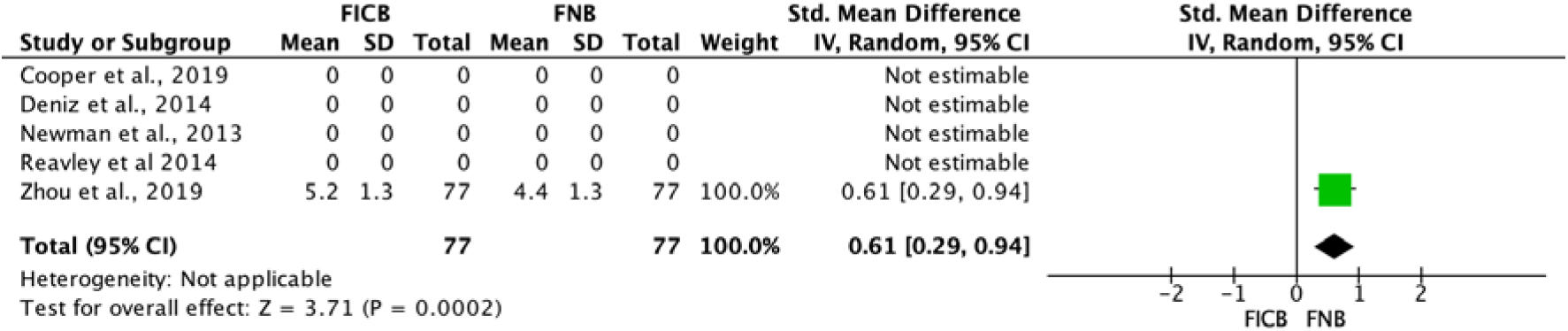
Forest plot of comparison: FICB vs. FNB. Outcome: pain scores on movement at 24 hours.

**Table 5.** Fascia iliaca compartment block vs. Femoral nerve block

| Outcome or Subgroup | Number of Studies | Participants | Statistical Method | Effect Estimate |
| --- | --- | --- | --- | --- |
| Initial Pain Scores at Rest Following Block | 5 | 523 | Std. Mean Difference (IV, Random, 95% CI) | 0.07 [-0.22, 0.35] |
| Pain Scores at Rest at 12 Hours | 5 | 0 | Std. Mean Difference (IV, Random, 95% CI) | Not estimable |
| Pain Scores at Rest at 24 Hours | 5 | 154 | Std. Mean Difference (IV, Random, 95% CI) | 0.10 [-0.22, 0.42] |
| Initial Pain Scores on Movement Following Block | 5 | 154 | Std. Mean Difference (IV, Random, 95% CI) | 0.53 [0.21, 0.86] |
| Pain Scores on Movement at 12 Hours | 5 | 0 | Std. Mean Difference (IV, Random, 95% CI) | Not estimable |
| Pain Scores on Movement at 24 Hours | 5 | 154 | Std. Mean Difference (IV, Random, 95% CI) | 0.61 [0.29, 0.94] |

Forest plots for the studies with statistically significant findings can be found in Figures 6-9.

**Figure 6.**
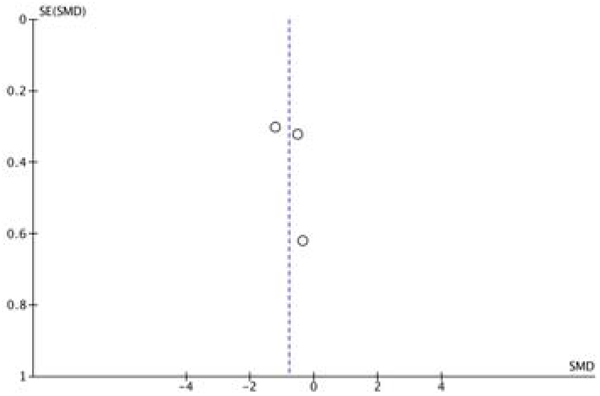
Funnel plot of comparison: FICB vs. Opioids. Outcome: pain scores at rest at 24 hours.

**Figure 7.**
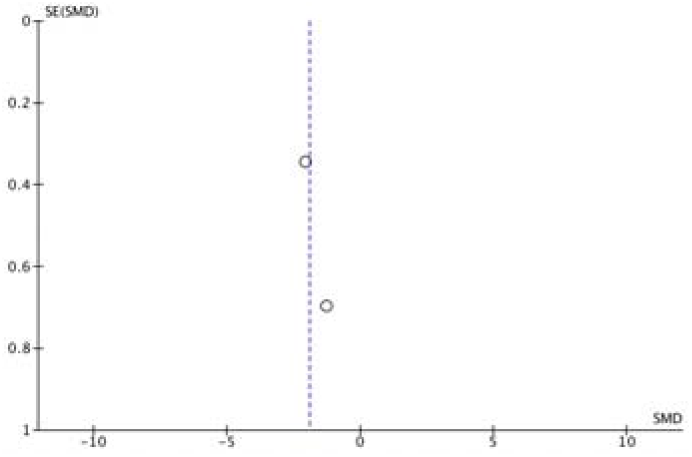
Funnel plot of comparison: FICB vs. Opioids. Outcome: pain scores on movement at 12 hours.

**Figure 8.**
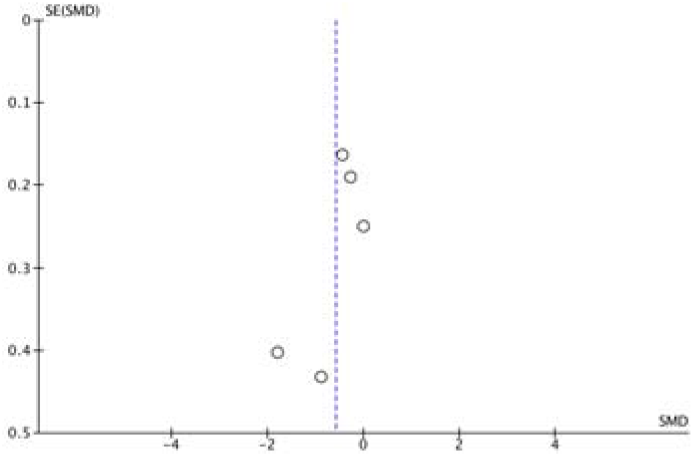
Funnel plot of comparison: FNB vs. Opioids. Outcome: initial pain scores at rest following block.

**Figure 9.**
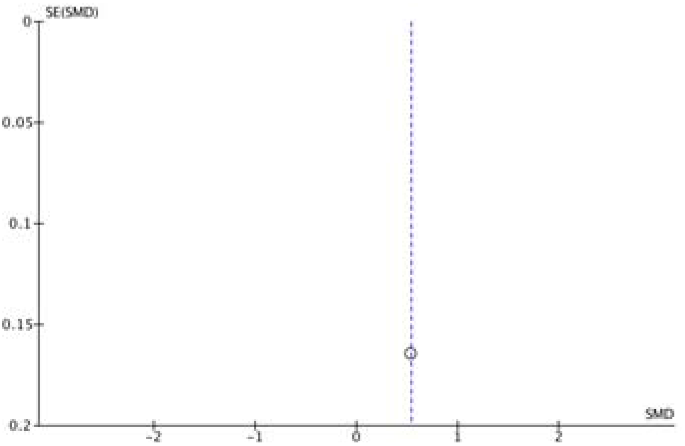
Funnel plot comparison: FICB vs. FNB. Outcome: initial pain scores on movement following block.

**Figure 10.**
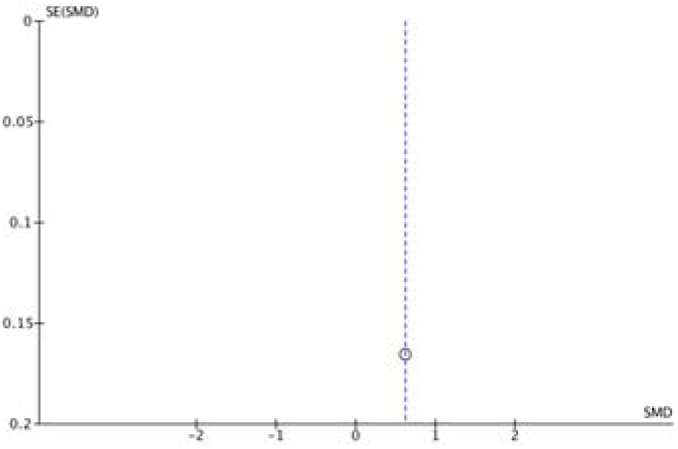
Funnel plot of comparison: FICB vs. FNB Outcome: pain scores on movement at 24 hours

## DISCUSSION

### Summary of main results

This systematic review and meta-analysis demonstrated that FNB had a superior analgesic effect compared with FICB, and both FNB and FICB had superior analgesic effect compared with opioids.

Of the twenty studies included in the meta-analysis, 7 studies compared FICB with opioids. Pain scores at rest at 24 hours (-0.79 [-1.34, -0.24], P= 0.005), and on movement at 12 hours post hip-fracture surgery (-1.91 [-2.5, -1.3], P< 0.00001) were lower in the FICB group compared with the opioid group. However, there was no statistically significant difference between the use of FICB versus opioids in analgesic efficacy measured by pain (VAS, NRS) scores after hip fracture surgery in patients initially at rest following the block, at rest at 12 hours, initial pain scores on movement following the block, or on movement at 24 hours.

An additional seven studies compared FNB with opioids. The initial pain scores at rest were lower in the FNB group (-0.58 [-0.104, -0.12], P= 0.01). However, there was no statistically significant difference between the use of FNB versus opioids in analgesic efficacy after hip fracture surgery in patients at rest at 12 hours, 24 hours, or when comparing initial pain scores on movement following the block. There was insufficient data for pain scores on movement at 12 hours, and 24 hours.

Finally, five studies compared FNB with FICB. Pain scores on movement initially following the block, and at 24 hours were lower in the FNB group when compared with FICB (initial: 0.53 [0.21, 0.86], P= 0.001, 24 h: 0.61 [0.29, 0.94], P= 0.0002). However, there was no statistically significant difference between the groups for pain scores at rest. There was insufficient data for pain scores at rest at 12 hours, and pain scores on movement at 12 hours.

The remaining study compared the analgesic efficacy of FICB with intra-articular hip injection (IAHI) as control[12]. The primary outcome was pain relief measured using the NRS at 20 minutes, 12 hours, 24 hours and 48 hours after the regional anaesthesia, both at rest and on movement. Pain was significantly lower in the IAHI group during movement of the fractured limb at 20 min (p < 0.05), 12h (p<0.05), 24h (p<0.05) and 48h (p<0.05). the findings of this study suggest that IAHI provides better pre-operative pain management in elder patients with intracapsular hip fractures compared to FICB.

### Strengths and Limitations

The strengths of this study are that two independent reviewers decided the eligibility of the full-text articles and conducted the risk of bias assessment, and high risk of bias papers were not included in the meta-analysis. In addition, a full manual search was conducted to ensure no missing articles, and authors of the included papers were contacted for missing data. However, the limitations of this study were that filters were applied during the search, which set limits for the type of study, age, language, etc. Due to subgrouping, there were not enough studies to conduct funnel plots and rule out publication bias. The funnel plot might appear symmetrical, but the interpretation is unsure, with only five to seven studies per subgroup. In addition, the statistically significant results for the FICB vs. FNB subgroup were based on data from 1 paper.

## AUTHORS’ CONCLUSIONS

### Implications for practice

Both femoral nerve block and fascia iliaca compartment block enhance analgesic outcomes following hip fracture and hip fracture surgery, superior to the use of systemic analgesics such as opioids. FNB may be more efficacious at reducing pain following hip fracture surgery when compared to FICB.

### Implications for research

This paper was designed to include all forms of regional anaesthesia for the lumbar plexus and its terminal branches. In the end, there was only enough high quality randomized-controlled trials to answer the research questions for Femoral nerve block and Fascia Iliaca Compartment block. More research should be conducted on the analgesic efficacy of PCB and PENG block for hip fracture surgeries. That way, future systematic reviews and meta-analyses can effectively compare all the block types. Future studies should further focus on comparing the analgesic efficacy of FNB vs FICB with respect to hip fracture surgeries to see if evidence supports the superiority of FNB compared to FICB.

## CONTRIBUTIONS OF AUTHORS

Conducted literature search and study selection: A.M.S.

Supervised literature search and study selection: B.O.D.

Performed data extraction and assessment of risk of bias: both authors.

Conducted meta-analysis: A.M.S.

Supervised meta-analysis: B.O.D.

Wrote manuscript: A.M.S.

## DIFFERENCES BETWEEN PROTOCOL AND REVIEW

There were no differences between the protocol and review.

## Data Availability

All data produced in the present work are contained in the manuscript

## ACKNOWLEDGMENT

I would like to thank Dr. Brian O’Donnell for his contributions and guidance throughout this project.

## APPENDIX

**Appendix A.**
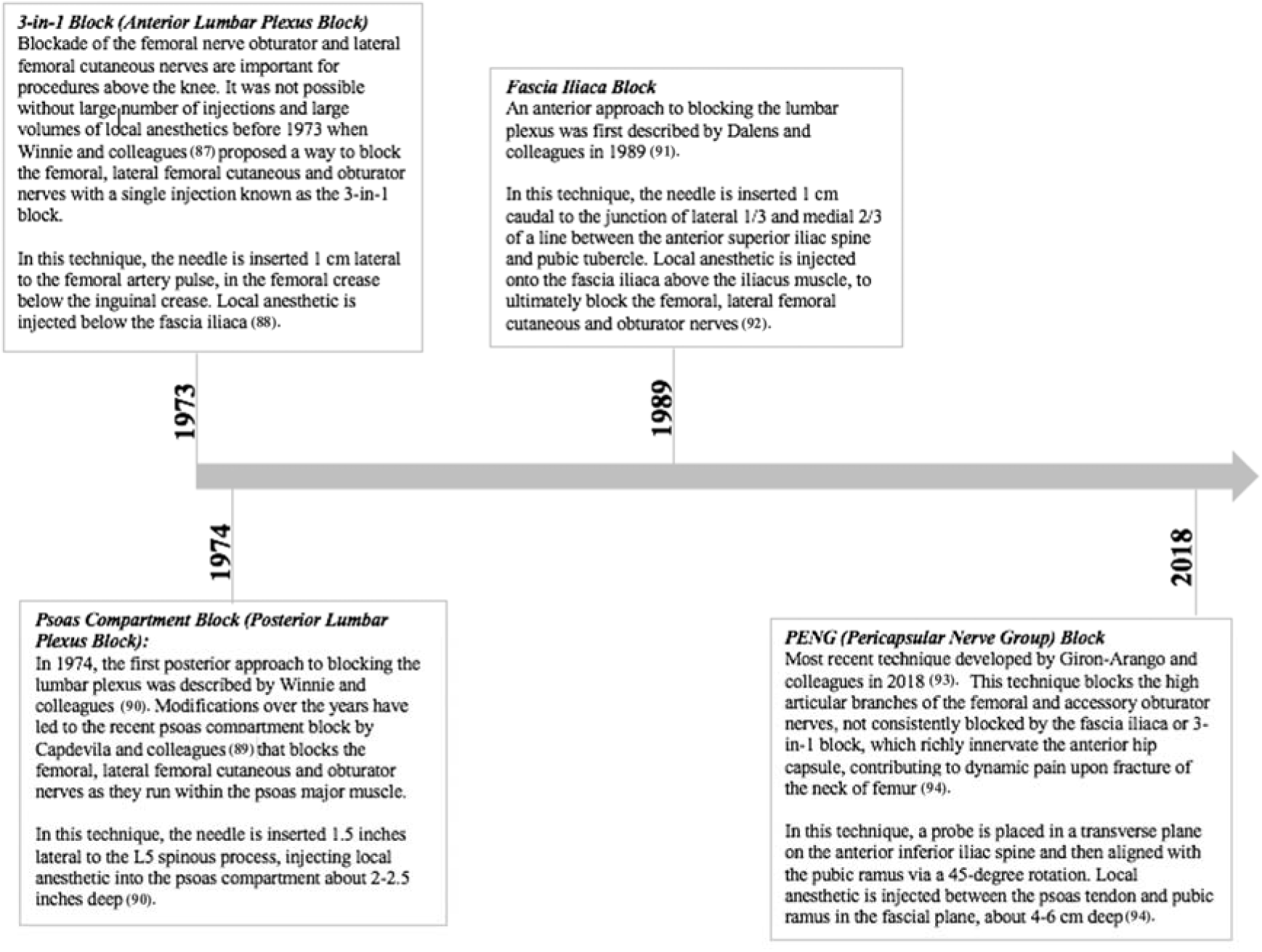
Timeline and description of regional anaesthesia approaches to the lumbar plexus and associated terminal branches.

**Appendix B.** Database search words and results.

| Database | Search Key | Number of Results |
| --- | --- | --- |
| EMBASE | ((('hip fracture'/exp OR 'hip fracture' OR (('hip'/exp OR hip) AND ('fracture'/exp OR fracture)) OR 'hip surgery'/exp OR 'hip surgery' OR (('hip'/exp OR hip) AND ('surgery'/exp OR surgery)) OR 'femur fracture'/exp OR 'femur fracture' OR (('femur'/exp OR femur) AND ('fracture'/exp OR fracture))) AND ('lumbar plexus block'/exp OR 'lumbar plexus block' OR (lumbar AND ('plexus'/exp OR plexus) AND block)) OR '3-in-1 block' OR ('3 in 1' AND block) OR 'peng block' OR (peng AND block) OR 'pericapsular nerve group block'/exp OR 'pericapsular nerve group block' OR (pericapsular AND ('nerve'/exp OR nerve) AND ('group'/exp OR group) AND block) OR 'fascia iliaca compartment block'/exp OR 'fascia iliaca compartment block' OR (('fascia'/exp OR fascia) AND iliaca AND compartment AND block) OR 'psoas compartment block'/exp OR 'psoas compartment block' OR (psoas AND compartment AND block)) AND 'randomized controlled trial'/de AND 'human'/de AND [english]/lim AND ([adult]/lim OR [aged]/lim OR [middle aged]/lim OR [very elderly]/lim) AND [article]/lim | 175 |
| PUBMED | (((((('hip fractures'[MeSH Terms] OR ('hip'[All Fields] AND "fractures"[All Fields]) OR "hip fractures"[All Fields] OR ('hip'[All Fields] AND "fracture"[All Fields]) OR "hip fracture"[All Fields]) OR (('hip'[MeSH Terms] OR "hip"[All Fields]) AND ("surgery"[Subheading] OR "surgery"[All Fields] OR "surgical procedures, operative"[MeSH Terms] OR ("surgical"[All Fields] AND "procedures"[All Fields] AND "operative"[All Fields]) OR "operative surgical procedures"[All Fields] OR "surgery"[All Fields] OR "general surgery"[MeSH Terms] OR ("general"[All Fields] AND "surgery"[All Fields]) OR "general surgery"[All Fields])) OR (('femoral fractures'[MeSH Terms] OR ("femoral"[All Fields] AND "fractures"[All Fields]) OR "femoral fractures"[All Fields] OR ("femur"[All Fields] AND "fracture"[All Fields]) OR "femur fracture"[All Fields]) AND ("lumbosacral plexus"[MeSH Terms] OR ("lumbosacral"[All Fields] AND "plexus"[All Fields]) OR "lumbosacral plexus"[All Fields] OR ("lumbar"[All Fields] AND "plexus"[All Fields]) OR "lumbar plexus"[All Fields]) AND block[All Fields])) AND ((3-in-1[All Fields] AND block[All Fields]) OR (PENG[All Fields] AND block[All Fields]) OR (Pericapsular[All Fields] AND Nerve[All Fields] AND ("population groups"[MeSH Terms] OR ("population"[All Fields] AND "groups"[All Fields]) OR "population groups"[All Fields] OR "group"[All Fields] AND Block[All Fields]) OR (('fascia'[MeSH Terms] OR "fascia"[All Fields]) AND iliaca[All Fields] AND compartment[All Fields] AND block[All Fields]) OR (Psoas[All Fields] AND compartment[All Fields] AND block[All Fields])))) AND (Randomized Controlled Trial[ptyp])) AND ("humans"[MeSH Terms])) AND (English[lang])) AND (('adult'[MeSH Terms] OR "adolescent"[MeSH Terms] OR "young adult"[MeSH Terms] OR "adult"[MeSH Terms:noexp] OR ("middle aged"[MeSH Terms] OR "aged"[MeSH Terms]) OR "middle aged"[MeSH Terms] OR "aged"[MeSH Terms] OR "aged, 80 and over"[MeSH Terms])) Filters: Randomized Controlled Trial, English | 57 |
| SCOPUS | TITLE-ABS-KEY (hip AND fracture AND nerve AND block) AND (LIMIT-TO (SRCTYPE, "j")) AND (LIMIT-TO (DOCTYPE, "ar" )) AND (LIMIT-TO (LANGUAGE, "English")) AND (LIMIT-TO (EXACTKEYWORD, "Randomized Controlled Trial")) | 45 |
| EBSCO:<br>CINAHL,<br>MEDLINE | (hip fracture AND nerve block) AND (RCT or randomized control trial or randomized controlled trial)<br>Limiters - Human<br>Expanders - Apply equivalent subjects<br>Narrow by Language: - English | 39 |
| WEB OF SCIENCE | TOPIC: (Hip fracture AND nerve block)<br>Refined by: DOCUMENT TYPES: (ARTICLE) AND TOPIC: (RCT OR randomized controlled trial OR randomized control trial) AND LANGUAGES: (ENGLISH) | 41 |
| COCHRANE LIBRARY | hip fracture AND nerve block in Title Abstract Keyword AND "randomized clinical trial" OR "randomized controlled trial" OR "randomized control trial" OR "RCT" in Title Abstract Keyword - (Word variations have been searched) | 82 |
| Total Articles: |  | 439 |

**Appendix C.** Inclusion and exclusion criteria used to select articles.

| Inclusion Criteria | Exclusion Criteria |
| --- | --- |
| Randomized controlled trials<br>Population<br>Scheduled for hip fracture surgery<br>Intervention<br>3-in-1 block<br>PENG block<br>Fascia iliaca compartment block (FICB)<br>Psoas compartment block (PCB)<br>Continuous catheter infusion<br>Single dose<br>Any dose or form of local anaesthetic to accompany technique<br>Comparison<br>Interventions of interest compared to each other<br>Placebo<br>Non-intervention<br>Opioids (any)<br>NSAIDs (any)<br>Paracetamol<br>Outcome: Analgesic Efficacy<br>Numeric Pain Rating Scale (NRS)<br>Visual Analogue Scale (VAS)<br>Pre-operative and/or post-operative need for analgesia<br>Time to first request for additional analgesia<br>Adverse effects<br>Allergic reactions<br>Length of hospital stay<br>Mortality | Articles unavailable in full text<br>Unavailable in English<br>Non-human studies<br>Include ages <18 years old<br>Conference proceedings<br>Systematic reviews<br>Narrative reviews<br>Meta-analyses<br>Surgery unrelated to hip |

**Appendix D.**
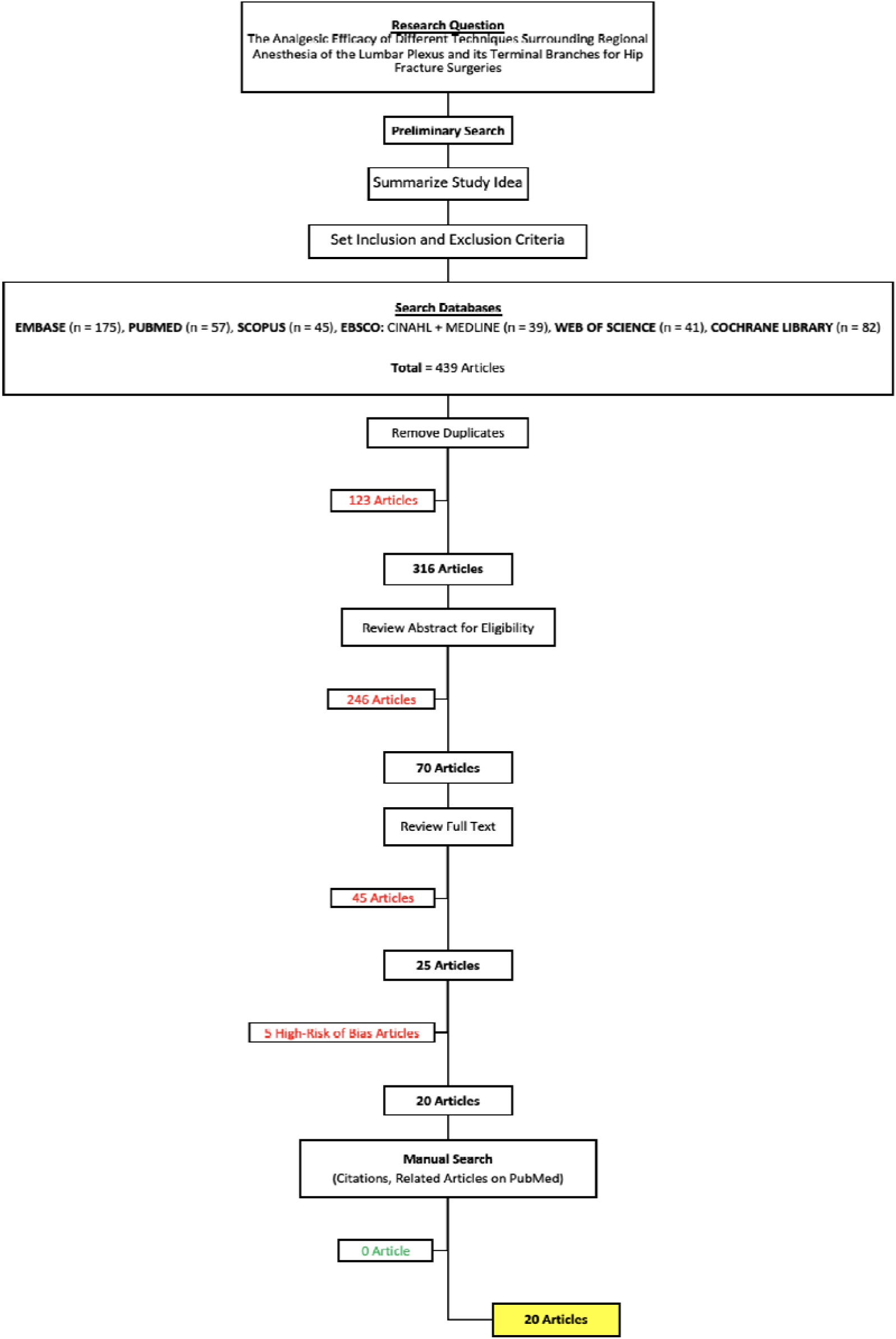
Overview of research protocol and study selection process.

**Appendix E.** completed checklist for the reporting guideline appropriate to study: PRISMA 2020 Checklist from www.equator-network.org

| Section and Topic | Item # | Checklist item |  |
| --- | --- | --- | --- |
| **TITLE** |  |  |  |
| Title | 1 | Identify the report as a systematic review. | X |
| **ABSTRACT** |  |  |  |
| Abstract | 2 | See the PRISMA 2020 for Abstracts checklist. | X |
| **INTRODUCTION** |  |  |  |
| Rationale | 3 | Describe the rationale for the review in the context of existing knowledge. | X |
| Objectives | 4 | Provide an explicit statement of the objective(s) or question(s) the review addresses. | X |
| **METHODS** |  |  |  |
| Eligibility criteria | 5 | Specify the inclusion and exclusion criteria for the review and how studies were grouped for the syntheses. | X |
| Information sources | 6 | Specify all databases, registers, websites, organisations, reference lists and other sources searched or consulted to identify studies. Specify the date when each source was last searched or consulted. | X |
| Search strategy | 7 | Present the full search strategies for all databases, registers and websites, including any filters and limits used. | X |
| Selection process | 8 | Specify the methods used to decide whether a study met the inclusion criteria of the review, including how many reviewers screened each record and each report retrieved, whether they worked independently, and if applicable, details of automation tools used in the process. | X |
| Data collection process | 9 | Specify the methods used to collect data from reports, including how many reviewers collected data from each report, whether they worked independently, any processes for obtaining or confirming data from study investigators, and if applicable, details of automation tools used in the process. | X |
| Data items | 10a | List and define all outcomes for which data were sought. Specify whether all results that were compatible with each outcome domain in each study were sought (e.g. for all measures, time points, analyses), and if not, the methods used to decide which results to collect. | X |
|  | 10b | List and define all other variables for which data were sought (e.g. participant and intervention characteristics, funding sources). Describe any assumptions made about any missing or unclear information. | X |
| Study risk of bias assessment | 11 | Specify the methods used to assess risk of bias in the included studies, including details of the tool(s) used, how many reviewers assessed each study and whether they worked independently, and if applicable, details of automation tools used in the process. | X |
| Effect measures | 12 | Specify for each outcome the effect measure(s) (e.g. risk ratio, mean difference) used in the synthesis or presentation of results. | X |
| Synthesis methods | 13a | Describe the processes used to decide which studies were eligible for each synthesis (e.g. tabulating the study intervention characteristics and comparing against the planned groups for each synthesis (item #5)). | X |
|  | 13b | Describe any methods required to prepare the data for presentation or synthesis, such as handling of missing summary statistics, or data conversions. | X |
|  | 13c | Describe any methods used to tabulate or visually display results of individual studies and syntheses. | X |
|  | 13d | Describe any methods used to synthesize results and provide a rationale for the choice(s). If meta-analysis was performed, describe the model(s), method(s) to identify the presence and extent of statistical heterogeneity, and software package(s) used. | X |
|  | 13e | Describe any methods used to explore possible causes of heterogeneity among study results (e.g. subgroup analysis, meta-regression). | X |
|  | 13f | Describe any sensitivity analyses conducted to assess robustness of the synthesized results. | X |
| Reporting bias assessment | 14 | Describe any methods used to assess risk of bias due to missing results in a synthesis (arising from reporting biases). | X |
| Certainty assessment | 15 | Describe any methods used to assess certainty (or confidence) in the body of evidence for an outcome. | X |
| **RESULTS** |  |  |  |
| Study selection | 16a | Describe the results of the search and selection process, from the number of records identified in the search to the number of studies included in the review, ideally using a flow diagram. | X |
|  | 16b | Cite studies that might appear to meet the inclusion criteria, but which were excluded, and explain why they were excluded. | X |
| Study characteristics | 17 | Cite each included study and present its characteristics. | X |
| Risk of bias in studies | 18 | Present assessments of risk of bias for each included study. | X |
| Results of individual studies | 19 | For all outcomes, present, for each study: (a) summary statistics for each group (where appropriate) and (b) an effect estimate and its precision (e.g. confidence/credible interval), ideally using structured tables or plots. | X |
| Results of syntheses | 20a | For each synthesis, briefly summarise the characteristics and risk of bias among contributing studies. | X |
|  | 20b | Present results of all statistical syntheses conducted. If meta-analysis was done, present for each the summary estimate and its precision (e.g. confidence/credible interval) and measures of statistical heterogeneity. If comparing groups, describe the direction of the effect. | X |
|  | 20c | Present results of all investigations of possible causes of heterogeneity among study results. | X |
|  | 20d | Present results of all sensitivity analyses conducted to assess the robustness of the synthesized results. | X |
| Reporting biases | 21 | Present assessments of risk of bias due to missing results (arising from reporting biases) for each synthesis assessed. | X |
| Certainty of evidence | 22 | Present assessments of certainty (or confidence) in the body of evidence for each outcome assessed. | X |
| **DISCUSSION** |  |  |  |
| Discussion | 23a | Provide a general interpretation of the results in the context of other evidence. | X |
|  | 23b | Discuss any limitations of the evidence included in the review. | X |
|  | 23c | Discuss any limitations of the review processes used. | X |
|  | 23d | Discuss implications of the results for practice, policy, and future research. | X |
| **OTHER INFORMATION** |  |  |  |
| Registration and protocol | 24a | Provide registration information for the review, including register name and registration number, or state that the review was not registered. | X |
|  | 24b | Indicate where the review protocol can be accessed, or state that a protocol was not prepared. | X |
|  | 24c | Describe and explain any amendments to information provided at registration or in the protocol. | X |
| Support | 25 | Describe sources of financial or non-financial support for the review, and the role of the funders or sponsors in the review. | X |
| Competing interests | 26 | Declare any competing interests of review authors. | X |
| Availability of data, code and other materials | 27 | Report which of the following are publicly available and where they can be found: template data collection forms; data extracted from included studies; data used for all analyses; analytic code; any other materials used in the review. | X |

